# Portable gynaecological imaging to enhance naked-eye visual assessment for the management of cervical precancer among women living with HIV in Kenya: An implementation-informed, diagnostic accuracy study protocol

**DOI:** 10.64898/2026.11.16.26368246

**Authors:** Vernon Mochache, Robert Osoti, Thomas Odeny, R. Scott McClelland

**Author notes:** ***Corresponding author:*** Dr. Vernon Mochache, University of Global Health Equity, P.O. Box 6955, Kigali, Rwanda.

## Abstract

**Background:** Cervical cancer remains a leading cause of morbidity and mortality in low- and middle-income countries, particularly among women living with HIV (WLHIV). Although visual-based assessment in primary care is feasible and facilitates same-day management, its diagnostic accuracy is dependent on provider experience and skill, resulting in variable performance. Portable gynecological imaging may enhance visual assessment while supporting implementation of cervical cancer prevention in primary care.

**Methods:** We will conduct a cross-sectional study among 150 WLHIV co-infected with human papillomavirus and their healthcare providers at Molo Sub-County Hospital in Nakuru County, Kenya. The study will evaluate the agreement between portable gynecological imaging-enhanced visual assessment and conventional colposcopy; estimate diagnostic accuracy using histopathology as the reference standard; determine provider-level acceptability, appropriateness, and feasibility, and assess patient-level satisfaction using validated instruments complemented by qualitative interviews and integrated using a sequential, explanatory, mixed-methods approach.

**Discussion:** This study will generate preliminary evidence on the diagnostic accuracy and implementation of portable gynecological imaging in primary care. If shown to be accurate, acceptable, appropriate, and feasible, portable gynecological imaging could improve timely identification and management of cervical precancer, and support expansion of high-quality cervical cancer prevention services in similar resource-limited settings.

## BACKGROUND

An estimated 600,000 new cases and almost 350,000 deaths as a result of invasive cervical cancer (ICC) occur worldwide each year. Of these, nearly 90% of new cases and more than 90% of deaths occur in low-and middle-income countries (LMICs)(1). Despite the increased life expectancy that has resulted from successful implementation of HIV prevention and care programs in these settings, the burden of ICC has remained disproportionately high among women living with HIV (WLHIV)(2). In nine LMICs with the highest HIV prevalence globally, the proportion of ICC cases attributable to HIV infection was ≥40% in 2020 (3). Conversely, in 122 countries with much lower HIV prevalence, this population attributable fraction was <5%. Compared with HIV-uninfected women, WLHIV are nearly twice as likely to be infected with high-risk human papillomavirus (HPV) subtypes, have higher rates of HPV persistence and recurrence after precancer management, and are six times more likely to progress to ICC (4, 5).

In 2020, the World Health Organization (WHO) launched the Global Strategy for Cervical Cancer Elimination (6). The strategy identifies WLHIV as a priority population and seeks to redirect global focus towards scaling up evidence-based interventions that respond to their needs. It has three pillars that include: i) enhancing primary prevention through HPV vaccination, ii) promoting secondary prevention through screening and early detection and, iii) timely management of precancer and ICC, or provision of palliative care for those with terminal disease. To complement this strategy, the WHO Guidelines for Screening and Treatment of Pre-Cancerous Lesions for the Prevention of Cervical Cancer (7) recommend that treatment decisions for cervical pre-cancer should be made based on either a positive primary screening test (i.e., *screen and treat*), or a positive primary screening and secondary triage tests (i.e., *screen-triage-treat*), with or without histological confirmation.

For primary screening, WHO recommends a high-performance HPV test for women between 25-50 years since it is associated with a higher sensitivity and samples can be self-collected, further improving acceptability (8–11). The higher sensitivity also allows for a longer screening interval for those who test negative (12), and the same sample can be used for genotyping to determine the presence of high-risk HPV subtypes (13–15). Among WLHIV, HPV-based primary screening has low to moderate specificity, meaning that a significant proportion of women who test HPV-positive do not eventually progress to ICC. In LMICs, the resultant unnecessary referrals and/or overtreatment further strain limited health systems (16, 17). For these reasons, WHO recommends a secondary triage test to allow for targeted management of women at high risk of progressing to precancer and/or ICC.

Visual assessment after application of either acetic acid (VIA) or Lugol’s iodine (VILI) has been widely used in LMICs. These procedures have low attendant costs and provide immediate results, eliminating the requirement for repeat health facility visits. Additionally, no specimen collection or specialized laboratory services are required (18). Visual assessment could be used for secondary triage with the added advantage of allowing assessment of eligibility for ablative treatment through visualization and characterization of the cervical transformation zone (TZ). Together with HPV-based primary screening, it possible to have a one-stop-shop approach (19, 20), increasing the feasibility, acceptability and appropriateness of implementing cervical cancer prevention in primary care. However, naked-eye visual assessment is user-dependent, with variable performance depending on the level of training and skill of the provider (21–23), and does not allow for a record of the screening encounter for quality improvement.

In recent years, emerging evidence suggests that portable gynecological imaging may perform similarly to conventional colposcopy in the identification of cervical pre-cancerous lesions (24–26). These portable devices could be deployed in primary care and used by any cadre of trained healthcare providers. Portable devices work by providing the additional illumination and magnification needed to better visualize and characterize the cervical TZ. In this way, portable gynecological imaging devices could potentially improve the accuracy of identifying pre-cancerous cervical lesions and assessment of eligibility for ablative treatment at primary care, allowing for immediate linkage to treatment. Additionally, these portable devices can be used to capture cervical images and/or videos which can act as a record of the screening encounter and/or be incorporated into an electronic medical records system to support clinical decision-making via either telemedicine or artificial intelligence/machine learning algorithms (27, 28).

## STUDY OBJECTIVES

The primary objective of this study is to generate preliminary evidence on diagnostic accuracy and implementation of using portable gynecological imaging to enhance naked-eye visual assessment in primary care. Specifically, we will determine the level of agreement between portable gynecological imaging compared to conventional colposcopy for the identification of cervical precancer and determining eligibility for ablative treatment. We will also determine the sensitivity, specificity, positive and negative predictive values (PPV and NPV) of identifying high-grade squamous intraepithelial lesions (HSIL+) using portable gynecological imaging. Finally, guided by Proctor’s Implementation Outcomes Framework, we will examine provider-level acceptability, appropriateness and feasibility and patient-level satisfaction with the proposed approach at primary care

## METHODS

### Study Setting

The study will be conducted at Molo Sub-County Hospital (SCH) in Nakuru County, Kenya. Molo SCH was one of the sites that participated in a Kenyan Ministry of Health (MoH) pilot study to assess HPV testing using the GeneXpert platform for cervical cancer screening between October 2019 and December 2020 where 328 (25%) of the 1,297 women targeted were screened, with a 24.4% HPV positivity rate (29). The health facility has both outpatient and inpatient services and provides HIV prevention, treatment, and care services to ~3,500 patients.

### Study Design

The proposed study will will adopt a cross-sectional design with procedures nested within ongoing clinical follow-up for WLHIV accessing prevention, treatment, and care services in Molo SCH.

### Study Population and Sample Size

A convenience sample of 150 HPV/HIV co-infected women aged 25–50 years old will be recruited for the diagnostic accuracy component of the study. This sample size will provide adequate precision for estimating the agreement between portable gynecological imaging-enhanced visual assessment and conventional colposcopy in identifying cervical precancer across an anticipated concordance range of 70%–90%. In addition, all healthcare providers directly involved in performing, supervising, or supporting the use of portable gynecological imaging within the HIV clinic (~8-10) will be invited to participate in the assessment of acceptability, appropriateness, and feasibility of integrating the technology into routine clinical practice.

### Eligibility

Women will be eligible if they receive HIV prevention, care, and treatment services at Molo SCH, are aged 25–50 years, test positive for HPV DNA, and provide written informed consent. They will be excluded if they decline or withdraw consent, are menstruating at the time of the scheduled cervical examination, are pregnant, have a history of total hysterectomy or previous cervical excisional treatment that would preclude valid colposcopic assessment. Healthcare providers will be eligible if they are currently employed at Molo SCH, are directly involved in performing, supervising, or supporting the use of portable gynecological imaging, and provide written informed consent. They will be excluded if they decline or withdraw consent, or if they have no experience using or supporting the implementation of the portable gynecological imaging device during the study period. Eligible providers may include nurses, clinical officers, medical officers, gynecologists, HIV clinicians, and other clinical personnel directly involved in cervical cancer screening, colposcopy, or implementation of study procedures.

### Study Procedures

We will mobilize potential study participants through an existing network of peer navigators. During routine HIV clinic visits, WLHIV will be screenend for inclusion in the study and requested to provide written informed consent. Those who consent will be asked to privately self-collect a vaginal specimen using the *Evalyn* cytobrush. We have previously demonstrated that Kenyan women preferred self-collection of genital specimens using this cytobrush (30) and their specimens perfomed similarly compared to clinician-collected samples for HPV testing (31). Participants will wait ~45 minutes for their HPV results from an on-site laboratory using the rapid, real-time, PCR-based Xpert HPV assay (Cepheid Inc., Sunnyvale, California, US).

WLHIV with a positive HPV DNA result will undergo VIA/VILI enhanced using a portable gynecological imaging device with characterization of their cervical TZ to determine eligibility for ablative treatment (7), and capture of cervical images and/or videos that will be automatically uploaded to an online, cloud-based repository. To maintain blinding and prevent bias during definitive diagnosis, the results of the enhanced visual assessment will not be provided to study participants. All HPV DNA positive participants, regardless of their enhanced visual assessment findings, will undergo definitive diagnosis using conventional colposcopy scheduled on the same day, where feasible and if not, within a week. To assess for verification bias, we will perform conventional colposcopy on 10% of WLHIV with HPV DNA negative screening results.

During conventional colposcopy, repeat visual assessment with determination of ablative treatment eligibility will be conducted. The colposcopist will collect at least four punch biopsies (i.e., one from each quadrant of the cervix, including biopsies of any visible lesions) for confirmatory histological assessment. Study participants will receive their visual assessment findings from this definitive diagnosis examination. In case the participant had a positive HPV DNA result but no visible cervical lesions, or if a visible lesion involves the cervical canal, the colposcopist will perform endocervical curettage (ECC). Punch-biopsy and ECC specimens will be sent to an external laboratory for histological assessment, with laboratory personnel blinded as to initial screening and triage findings.

All study participants determined to be eligible for ablative treatment from the conventional colposcopy examination will be offered either cryotherapy or thermal ablation on the same study visit, where feasible. Ineligible participants will be advised to return within a week for their histology results. Those with cervical intraepithelial neoplasia grade 2 and above (CIN2+) will undergo excisional treatment using either loop electro-excision procedure (LEEP) or large loop excision of the transformation zone (LLETZ). Participants with a pre-cancerous endocervical lesion or histologically-confirmed ICC will be referred to the Nakuru County Comprehensive Cancer Center for further management that may include cold knife conization, hysterectomy, chemotherapy and/or radiotherapy. Histology specimens will be archived for external quality control (EQC) and potential future testing. Study participants will receive reimbursement for transportation and other obligations related to the research study.

We will use a sequential, explanatory mixed-methods approach (32) to assess the implementation of portable gynecological imaging within routine HIV primary care from both the primary care provider and WLHIV perspectives. Quantitative data will be collected first to evaluate provider-level implementation outcomes at least one month after study initiation using the validated Acceptability of Intervention Measure (AIM), Intervention Appropriateness Measure (IAM), and Feasibility of Intervention Measure (FIM)(33). Patient satisfaction with the integrated service delivery model will be assessed immediately following the clinical encounter using the validated 8-item Client Satisfaction Questionnaire (CSQ-8), supplemented by additional questions exploring perceived privacy, procedural comfort, and the contribution of the portable gynecological imaging device to the overall care experience.

Following analysis of the quantitative findings, qualitative data will be collected through in-depth interviews (IDIs) with a purposively selected sample of WLHIV and key informant interviews (KIIs) with healthcare providers directly involved in implementation of the portable gynecological imaging. The qualitative inquiry will be informed by the quantitative results and designed to explain, elaborate, and contextualize observed implementation outcomes and patient experiences. Patient interviews will explore experiences with the integrated screening service, while provider interviews, guided by the Consolidated Framework for Implementation Research (CFIR), will examine determinants influencing adoption, workflow integration, training adequacy, implementation climate, and sustainability. Findings from both phases will be integrated during interpretation to explain quantitative results, identify factors influencing implementation, and inform refinement of the intervention and the design of a future hybrid effectiveness-implementation trial.

## DATA MANAGEMENT

### Data Collection

Quantitative data, including participant demographic and clinical characteristics, diagnostic assessment findings, provider implementation outcome measures, and patient satisfaction questionnaires, will be collected electronically using structured questionnaires developed on the KoboCollect platform and administered on password-protected smartphones or tablets. Qualitative data will be collected through audio-recorded IDIs/KIIs conducted by trained qualitative researchers using semi-structured interview guides in the participant’s preferred language (English or Swahili). Interview recordings will be supplemented with field notes, transcribed verbatim, translated into English where necessary, and de-identified before analysis.

All study data will be linked using the unique 10-digit participant identification code assigned through the Kenya national HIV program. This identifier will facilitate linkage of study questionnaires, clinical and laboratory data, cervical images, and qualitative interview transcripts while maintaining participant confidentiality. No directly identifiable personal information, including participant names or national identification numbers, will be entered into the study database. Electronic study data, interview transcripts, and audio recordings will be stored on secure, password-protected computers with access restricted to authorized study personnel. Audio recordings will be retained only until transcription and quality assurance have been completed, after which they will be permanently deleted in accordance with institutional data management procedures.

All personal data will be managed in accordance with the Kenya Data Protection Act (2019) and institutional data governance policies. Data will be pseudonymized wherever possible, and any sharing of study data with collaborators, sponsors, or regulatory authorities will occur only with participants’ informed consent, for purposes specified in the approved study protocol, and under formal data-sharing agreements. Appropriate technical and organizational safeguards, including encrypted data transfer, role-based access controls, and secure storage, will be implemented to protect participant confidentiality and prevent unauthorized access, disclosure, loss, or misuse of study data.

### Data Analysis

Participant demographic, clinical, obstetric, and behavioral characteristics will be summarized using frequencies and percentages for categorical variables and medians with interquartile ranges (IQRs) for continuous variables, as appropriate. Statistical analyses will be performed using Stata version 18 (StataCorp LLC, College Station, TX, USA), with statistical significance assessed at a two-sided α level of 0.05. For Specific Aim 1, agreement between portable gynecological imaging-enhanced visual assessment and conventional colposcopy will be assessed using Cohen’s kappa statistic. For Specific Aim 2, diagnostic accuracy will be evaluated using histopathology as the reference standard, with two-by-two contingency tables constructed to estimate sensitivity, specificity, PPV, and NPV together with their corresponding 95% confidence intervals.

Quantitative implementation outcomes will be measured using AIM/IAM/FIM, respectively. Patient satisfaction will be summarized using total CSQ-8 scores, while responses to supplementary questions on privacy, procedural comfort, and perceived contribution of the portable gynecological imaging device will be summarized descriptively. Qualitative data obtained from IDIs and KIIs will be analyzed using thematic content analysis. De-identified transcripts will be imported into either NVivo (QSR International) or ATLAS.ti (Scientific Software Development GmbH) to facilitate data management and coding. Two members of the research team will independently review and code the transcripts before developing an agreed coding framework. Coding will be both deductive, informed by CFIR constructs, and inductive to allow the identification of emergent themes.

Consistent with the sequential, explanatory mixed-methods design, qualitative findings will be used to explain, elaborate, and contextualize the quantitative implementation outcomes and patient satisfaction results. Integration will occur during interpretation through comparison of quantitative and qualitative findings to identify areas of convergence, complementarity, and divergence. The integrated findings will provide a comprehensive understanding of the factors influencing implementation of portable gynecological imaging within HIV primary care and will inform refinement of the intervention and the design of a future hybrid effectiveness-implementation trial.

## ETHICAL CONSIDERATIONS

This study will be conducted in accordance with the ethical principles of the Declaration of Helsinki, Good Clinical Practice (GCP) and Good Clinical Laboratory Practice (GCLP) guidelines, as well as applicable national regulatory requirements. Ethical approval has been obtained from the Kenya Medical Research Institute (KEMRI) Scientific and Ethics Review Unit

(SERU) (No. 5180) and the Institutional Review Board of the University of Washington (UW) Human Subjects Division (HSD)(STUDY00022950). Approval to conduct the study has also been obtained from the Kenya National Commission for Science, Technology and Innovation (NACOSTI/P/25/4176060) and the Nakuru County Department of Health Research Unit. All study personnel will complete GCP and Human Research Participants Protection training before study initiation, while laboratory personnel will complete (GCLP) training. Any protocol modification that may affect the scientific integrity of the study, participant safety, study objectives, study design, eligibility criteria, or study procedures will require a formal protocol amendment submitted to KEMRI SERU and UW HSD for ethical review and approval before implementation.

## DISSEMINATION

Study findings will be disseminated through stakeholder engagement meetings with the Ministry of Health, Nakuru County Department of Health, participating health facilities, community representatives, and other relevant stakeholders. Results will also be presented at national and international scientific conferences and published in peer-reviewed journals. Policy briefs will be developed to support translation of the findings into cervical cancer prevention policy and practice in Kenya and other LMICs. This study has been designed with consideration of established methodological and reporting standards. Findings from the diagnostic accuracy component will be reported in accordance with the Standards for Reporting Diagnostic Accuracy Studies (STARD 2015), the qualitative findings will be reported using the Consolidated Criteria for Reporting Qualitative Research (COREQ), and the mixed-methods findings will be reported in accordance with the Good Reporting of A Mixed Methods Study (GRAMMS) recommendations.

## Data Availability

No datasets have been generated or analysed for the current study. All relevant data from this study will be made available upon study completion.

## DECLARATIONS

### Human Ethics and Consent to Participate

Ethical approval for this study has been obtained from KEMRI SERU (No. 5180) and UW HSD (STUDY00022950). A research permit has been obtained from NACOSTI (NACOSTI/P/25/4176060) and the Nakuru County Department of Health Research Unit. The study will be conducted in accordance with the ethical principles of the Declaration of Helsinki, GCP) and GCLP guidelines, as well as applicable national regulatory requirements. All participants will provide written informed consent.

### Author Contributions

VM & RSM conceptualized the study, sought funding and prepared the first draft of this manuscript. RO, TO and SM contributed to development of the protocol and reviewed this manuscript for proper intellectual content. All authors read and approved the final draft of this manuscript.

## Acknowledgements

The authors would like to acknowledge the Nakuru County Department of Health, Molo SCH management as well as Cerba Lancet Laboratories.

## SUPPLEMENTARY FILE

Data collection tools and informed consent forms

